# Breakthrough Covid-19 infections during periods of circulating Beta, Delta and Omicron variants of concern, among health care workers in the Sisonke Ad26.COV2.S vaccine trial, South Africa

**DOI:** 10.1101/2021.12.21.21268171

**Authors:** Ameena Goga, Linda-Gail Bekker, Nigel Garrett, Tarylee Reddy, Nonhlanhla Yende-Zuma, Lara Fairall, Harry Moultrie, Azwidihwi Takalani, Valentina Trivella, Mark Faesen, Veronique Bailey, Ishen Seocharan, Glenda E Gray, on behalf of the Sisonke Study Team

## Abstract

**Background:** We report breakthrough infections (BTIs) during periods of circulating Beta, Delta and Omicron variants of concern, among health care workers (HCW) participating in the Sisonke phase 3B Ad26.COV2.S vaccine trial (ClinicalTrials.gov number, NCT04838795). Data were gathered between 17 February and 15 December 2021. Duration of each period in this study was 89 days for Beta, 180 days or Delta and 30 days for Omicron.

**Results:** A total of 40 538 BTIs were observed, with 609 during Beta, 22 279 during Delta and 17 650 during Omicron. By 15 December, daily infections during Omicron were three times that seen during the peak observed during Delta. However, unlike the Delta period, with Omicron there was a clear and early de-coupling of hospitalisation from cases as a percentage of the Delta peak curves. Omicron significantly infected a greater proportion of HCW in the 18-30 year age-group, compared with the 55+ age group. There were 1 914 BTI-related hospitalisations - 77, 1 429 and 408 in the Beta (89 days), Delta (180 days) and Omicron (30 days) periods, respectively. During Omicron, 91% hospitalized HCWs required general ward care, 6% high care and 3% intensive care, compared with 89% general ward care, 4% high care and 7% intensive care, during Delta and 78% general care, 7% high care and 16% intensive care during Beta (p<0.001). During Beta and Beta 43% of hospitalized HCW needed supplementary oxygen and 7-8% needed ventilation, compared with 16% and 0.2% respectively during the Omicron period (p<0.001). Median length of hospitalization was significantly lower with Omicron compared with Beta and Delta (3 days compared with 5-6 days, p<0.001).

**Conclusions:** We illustrate more BTIs but reassuringly less severe Covid-19 with Omicron. Re-infections and Omicron-driven primary infections were likely driven by high population SARS-CoV-2 seroprevalence, waning vaccine effectiveness over time, increased Omicron infectivity, Omicron immune evasion or a combination of these and need further investigation. Follow-up of this cohort will continue and reports will be updated, as time and infections accrue.

## Introduction

South Africa reported a new SARS-CoV-2 variant in November 2021.^1-3^ This variant, named Omicron, declared a variant of concern (VOC) on 26 November 2021^4^, spread exponentially, replacing Delta, and driving rapid increases in Covid-19 cases.^5,6^ In vitro experiments demonstrate that Omicron escapes antibody neutralization in previously infected or vaccinated.^7,8^ Epidemiological data suggest reduced vaccine effectiveness^9^ and higher re-infection rates compared with Beta and Delta VOC.^10^ There is sparsity of data on the severity of Omicron-driven break-through infections (BTI), defined as positive SARS-CoV-2 polymerase chain reaction or antigen tests 28 days or more post vaccination. We describe BTIs during periods of circulating Beta, Delta and Omicron variants of concern, among health care workers (HCW) participating in the Sisonke phase 3B Ad26.COV2.S vaccine trial (ClinicalTrials.gov number, NCT04838795).^11^ The Sisonke trial was conducted in up to 350 vaccination centres, across all nine provinces of South Africa. Study procedures included an electronic consent process, on-site check for vaccination eligibility, and post-vaccination safety monitoring. Using the national Electronic Vaccination Data System (EVDS), HCW self-reported demographic characteristics and comorbidities and vaccinators recorded vaccination details. The trial administered a single dose Ad26.COV2.S vaccine to 477 234 HCWs between 17 February and 17 May 2021; 230 488 HCW voluntarily received a second Ad26.COV2.S dose between 9 November and 16 December 2021. We evaluate BTI frequency and severity between the 3 March and 15 December 2021, using proxy dates for the three VOC periods: Beta (17 February (Sisonke study start)-17 May 2021 (89 days)), Delta (18 May-14 November 2021 (180 days)) and Omicron (15 November-15th December 2021 (30 days)).

## Methods

BTIs were monitored using active and passive surveillance. Linkage of Sisonke trial data with the Covid-19 Notifiable Medical Conditions Sentinel Surveillance (NMCSS) master list, DATCOV (COVID-19 related hospitalizations) list and National Population Register held by the South African Medical Research Council identified participants with Covid-19 infections/re-infections, Covid-19-related hospitalisations and deaths. Additionally, Sisonke trial participants received text messages post-vaccination encouraging them to report BTIs telephonically or via a web-link. The protocol safety team contacted all hospitalised participants to confirm severity and ascertain outcome. HCWs with re-infections were included in all relevant study periods.

## Results

A total of 40 538 BTIs were observed, with 609 during Beta, 22 279 during Delta and 17 650 during Omicron. By 15 December, daily infections during Omicron were three times that seen during the peak observed during Delta (Supplementary Figure 1). However, unlike the Delta period, with Omicron there was a clear and early de-coupling of hospitalisation from cases as a percentage of the Delta peak curves (Figure 1).

**Figure 1:**
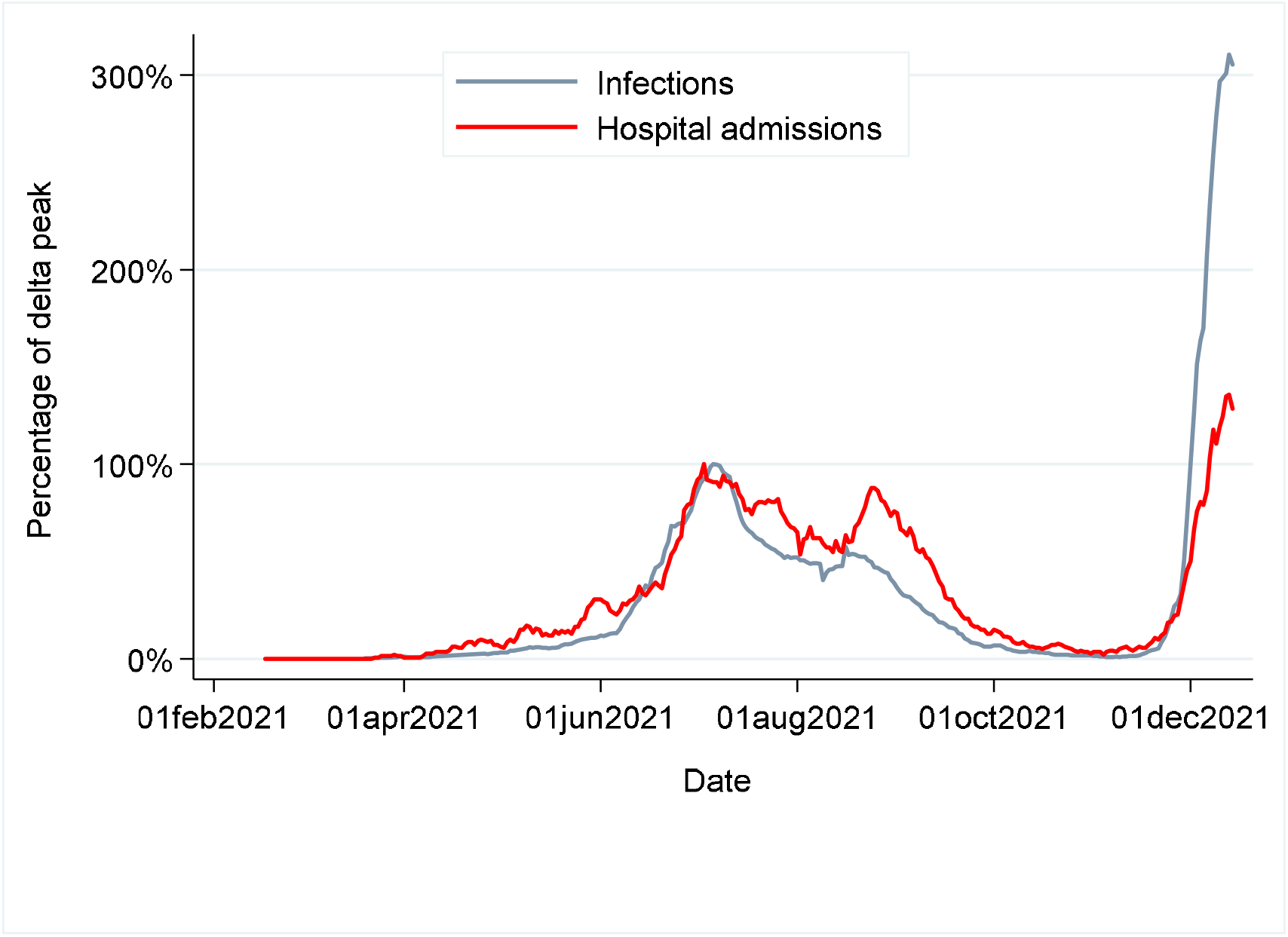
Daily cases and admissions expressed as a proportion of delta peak.

Omicron significantly infected a greater proportion of HCW in the 18-30 year age-group - 15% during Delta versus 21% during Omicron, and fewer in the 55+ age group (20% during Delta versus 13% during Omicron, p<0.001), Table 1.

**Table 1:**
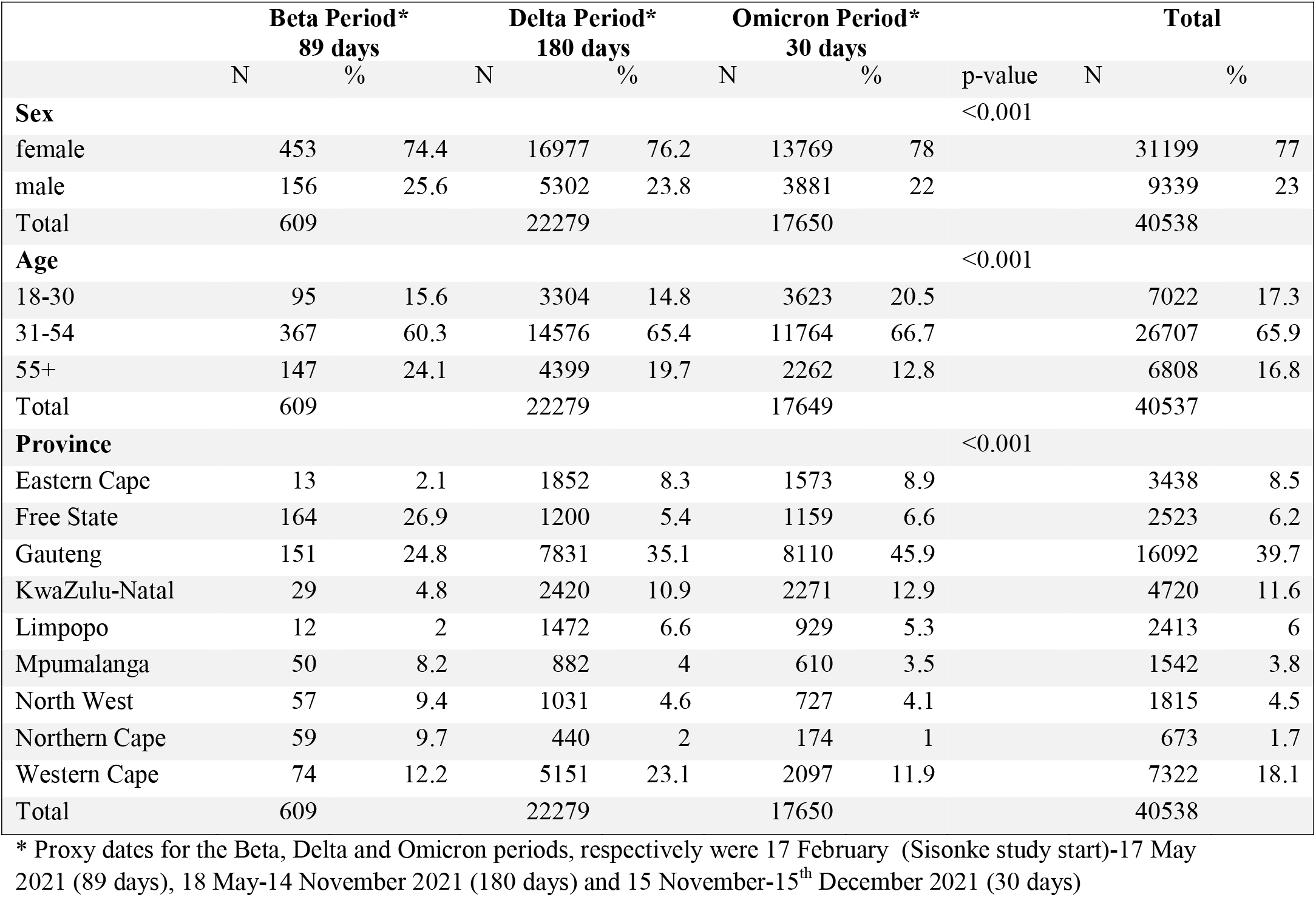
Description of breakthrough infections amongst vaccinated HCW for the Beta, Delta and Omicron periods, respectively.

There were 1 914 BTI-related hospitalisations - 77, 1 429 and 408 in the Beta, Delta and Omicron periods, respectively (Table 2). The proportion of BTI-related hospitalisations in the 31-54 year age group significantly increased from 51% during Beta to 60% during Delta and 67% during the Omicron periods, with a concomitant reduction in the 55+ age group (46% during Beta, to 33% during Delta and 19% during Omicron (p<0.001; Table 2)). Compared with the Beta and Delta periods, the prevalence of hypertension and diabetes in hospitalized HCW was significantly lower during Omicron, (Table 2); 47% hypertension and 25% diabetes during Beta, respectively, versus 35% and 23% during Delta and 23% and 10% respectively during Omicron, p<0.001.

**Table 2:**
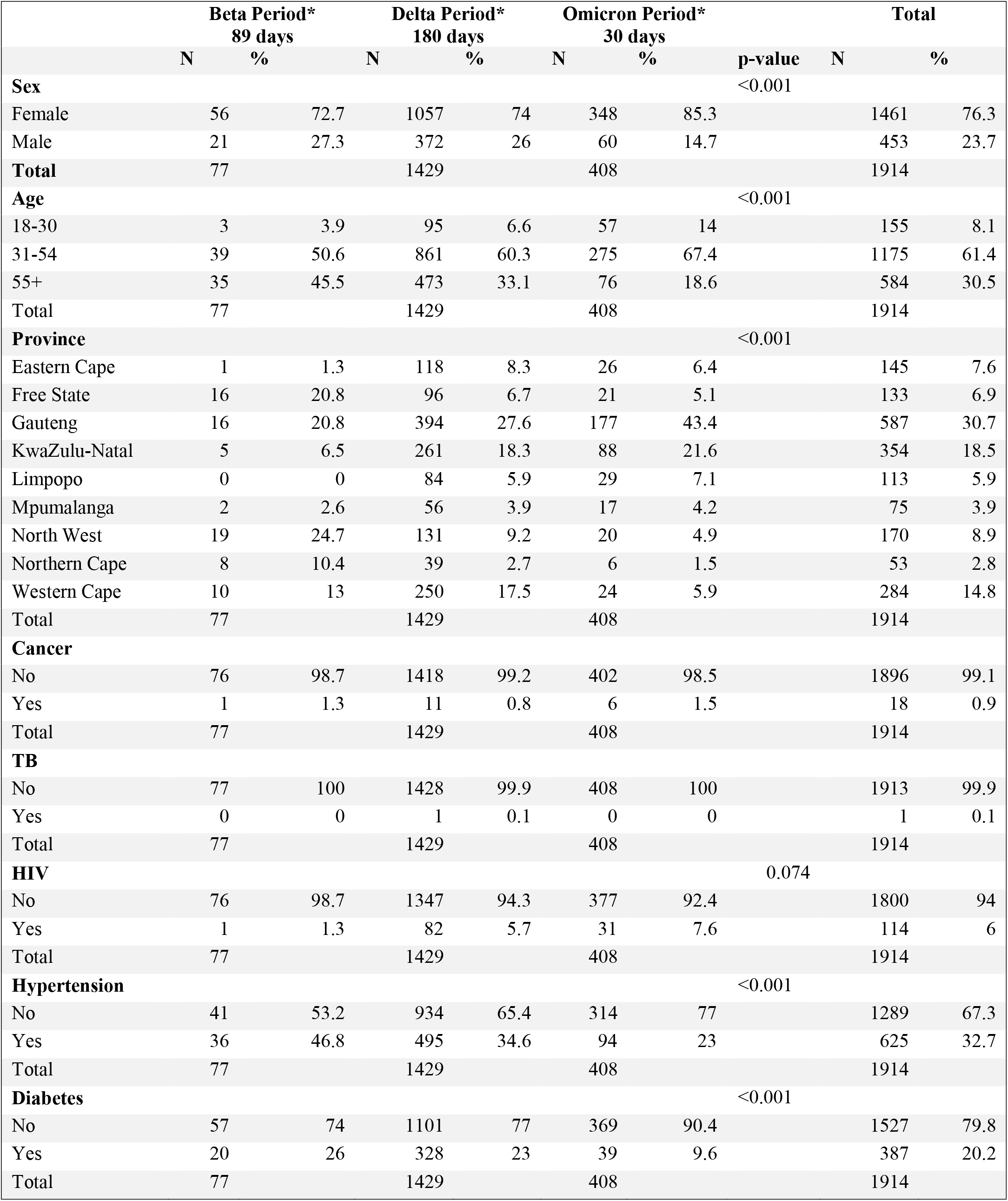
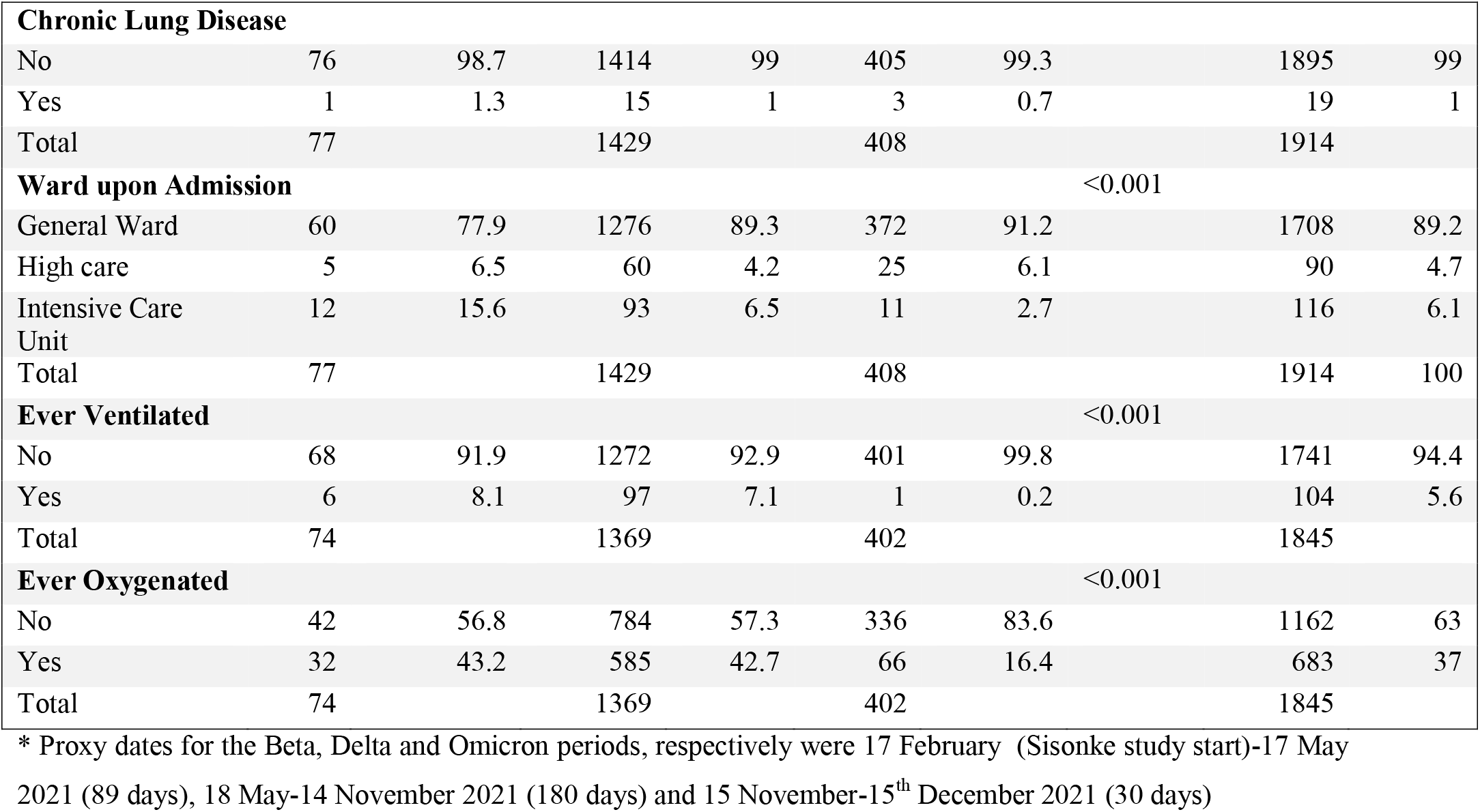
Description of characteristics of hospitalized, vaccinated HCW by period.

During Omicron, 91% hospitalized HCWs required general ward care, 6% high care and 3% intensive care. This was significantly different from 89% general ward care, 4% high care and 7% intensive care, during Delta and 78% general care, 7% high care and 16% intensive care during Beta (p<0.001).

During Beta and Beta 43% of hospitalized HCW needed supplementary oxygen and 7-8% needed ventilation, compared with 16% and 0.2% respectively during the Omicron period (p<0.001).

Amongst HCWs discharged from hospital with available data (n=1 780 of 1914), the median length of hospitalisation was 6 days (IQR 4-11; n=77) for Beta, 5 days (IQR 3-9; n=1416) for Delta and 3 days (IQR 1-5; n=287) days for Omicron, with significant differences between Omicron and Delta, p<0.0001.

Of the 17 650 infections during the Omicron period, 28 (2%) were previously infected during the Beta period and 786 (5%) during the Delta period, possibly signifying some cross-protection from Beta VOC infections. There were no significant differences in baseline characteristics (age, sex, comorbidities) between re-infected HCW who were initially infected during the Beta versus Delta periods.

## Discussion

We report on the first 30-days of the Omicron period and it may be too early to measure the full effect of the Omicron VOC. However, 30 days into the Omicron period, BTI cases had far surpassed that seen during the Delta peak. We did not account for person-time in follow-up - we present proportions by period. We included all hospitalized BTIs across all study periods, including incidental diagnoses amongst asymptomatic HCWs hospitalized for other reasons e.g. surgery; thus the number and proportion of BTIs needing Covid-19-related hospitalisation is likely over-estimated. Lastly, the course of COVID-19 due to Omicron in South Africa may be tempered by high population SARS-CoV-2 seroprevalence which was as high as 68% in some populations by April 2021; thus our findings may not be generalizable to all settings or populations globally.^12^

Despite these limitations, our large dataset provides an early snapshot of the effect of Omicron in a low-middle income, high SARS-CoV-2 seroprevalence setting. We illustrate more BTIs but reassuringly less severe Covid-19 with Omicron. Re-infections and Omicron-driven primary infections were likely driven by high population SARS-CoV-2 seroprevalence, waning vaccine effectiveness over time, increased Omicron infectivity, Omicron immune evasion or a combination of these and need further investigation. Follow-up of this cohort will continue and reports will be updated, as time and infections accrue.

## Data Availability

All data produced in the present study are available upon reasonable request to the authors

